# Statistical inference for association studies using electronic health records: handling both selection bias and outcome misclassification

**DOI:** 10.1101/2019.12.26.19015859

**Authors:** Lauren J. Beesley, Bhramar Mukherjee

## Abstract

Health research using electronic health records (EHR) has gained popularity, but misclassification of EHR-derived disease status and lack of representativeness of the study sample can result in substantial bias in effect estimates and can impact power and type I error. In this paper, we develop new strategies for handling disease status misclassification and selection bias in EHR-based association studies. We first focus on each type of bias separately. For misclassification, we propose three novel likelihood-based bias correction strategies. A distinguishing feature of the EHR setting is that misclassification may be *related to patient-specific factors*, and the proposed methods leverage data in the EHR to estimate misclassification rates *without gold standard labels*. For addressing selection bias, we describe how calibration and inverse probability weighting methods from the survey sampling literature can be extended and applied to the EHR setting.

Addressing misclassification and selection biases simultaneously is a more challenging problem than dealing with each on its own, and we propose several new strategies to address this situation. For all methods proposed, we derive valid standard errors and provide software for implementation. We provide a new suite of statistical estimation and inference strategies for addressing misclassification and selection bias simultaneously that is tailored to problems arising in EHR data analysis. We apply these methods to data from The Michigan Genomics Initiative (MGI), a longitudinal EHR-linked biorepository.

## 1 Introduction

Health research using data from large observational databases such as electronic health records (EHR) has gained popularity, and interest in such analyses continues to increase (Beesley et al., 2018b; Wolford et al., 2018). Unlike curated and well-designed population-based studies, these databases are rarely originally intended for research use and, consequently, patient recruitment processes may not be well understood (Casey et al., 2016). Without properly accounting for these design issues (e.g. who is in the study sample, how data were measured), association analyses using these data are naturally susceptible to bias (Beesley et al., 2018a). With larger and larger datasets at researchers’ fingertips, the impact of bias relative to variance is becoming more and more pronounced. In particular, these biases do not disappear with increased sample size, but the variance of the estimates does decrease with sample size, resulting in a greater potential for “incorrect” inference with inflated type 1 error and suboptimal coverage. This phenomenon is known as the “big data paradox” (Meng et al., 2018), and statistical strategies for correcting these biases are needed. In this paper, we focus on two common sources of bias for EHR data analysis: (1) misclassification of derived disease status (information bias) and (2) lack of representativeness (selection bias). We consider a very common inference problem where one is interested in relating a binary disease phenotype *D* with predictors *Z*.

EHR-derived disease status variables (phenotypes) can be misclassified for many reasons. Researchers often define disease status based on diagnosis codes recorded in the EHR for billing purposes, which provide a restricted snapshot of a patient’s complete disease history. Even the most sophisticated phenotyping methods are limited by the information available in structured and unstructured content of the EHR (e.g. Liao et al., 2015). Secondary conditions may not always be recorded, past medical history may be incomplete, and symptoms between visits may be missed. For academic databases, patients may visit the hospital for short-term treatment and return to local providers for continued care. These factors can lead to a large degree of mis-classification, particularly due to underreporting of disease. Several researchers have explored misclassification in EHR or claims data assuming *constant* sensitivity and specificity (Sinnott et al., 2014; Lange et al., 2015; Hubbard et al., 2015). However, a key feature of misclassification for EHR-derived phenotypes is that we expect more diagnoses to be missed for patients followed for a shorter period of time and for fewer visits, so *misclassification may depend on patients’ individual characteristics*. This problem has been discussed in the literature on HER data analysis (e.g. Bower et al., 2017; Goldstein et al., 2016; Phelan et al., 2017). Even so, statistical literature handling this covariate-related misclassification is sparse. Neuhaus (1999) presented analytic expressions for bias under covariate-related misclassification, and Beesley et al. (2018a) provided a sensitivity analysis approach tailored to the EHR setting. Ad hoc strategies including adjusting for number of encounters or clinic type have also been proposed (Goldstein et al., 2016; Phelan et al., 2017). In general, however, existing work considering covariate-related misclassification is limited, necessitating new statistical methods that can address this more complex misclassification mechanism.

EHR data are also susceptible to bias due to a lack of representativeness of the study sample with respect to some population of interest, e.g. the US population. Interactions with the health care system are generated by the patient, as is consent for biobank inclusion. Thus, it can be difficult to understand the mechanism driving patient inclusion, which may be related to a broad spectrum of patient factors including overall health and access to care. When ignored, patient selection related to disease status can often have a large impact on results of association analyses (Beesley et al., 2018b). Complex patient selection can often be addressed using survey techniques if the selection strategy is known, but a primary challenge for addressing patient selection in EHR is that the mechanisms for patient selection/inclusion are *unknown*. Researchers have attempted to partially account for selection bias by adjusting for patient factors such as referral status and clinic type (Phelan et al., 2017; Goldstein et al., 2016). Haneuse and Daniels (2016) developed a statistical framework for modeling selection in EHR data as a series of selection steps. This strategy can be very useful for characterizing selection mechanisms generating an analytical sample (e.g. patients aged 60+ with at least 6 months of follow-up) from a bigger EHR database. However, these methods do not address the systematic differences between patients in the population that are and are not included in the EHR itself. To bridge this gap, strategies in the survey sampling literature for dealing with unknown selection probabilities (termed non-probability sampling) such as calibration weighting, inverse probability of selection weighting, and propensity-score matching or covariate adjustment can be applied (Bower et al., 2017; Baker et al., 2013). Little work has been done to describe how such methods can be implemented in the specific EHR setting.

In this paper, we develop new, practical strategies for handling phenotype misclassification and selection bias in EHR-based association studies. We first focus on each type of bias separately. For misclassification, we propose three novel likelihood-based bias correction and inference strategies. These strategies allow us to estimate the rate of misclassification incorporating covariate relationships and require minimal external information and *no gold standard labels*. For addressing selection bias without misclassification, we describe how calibration and inverse probability of selection weighting methods from the survey sampling literature can be modified and applied in the EHR setting.

Addressing both sources of bias at once is a more challenging problem than dealing with each source of bias on its own, and we propose several new estimation and inference strategies. For all bias correction strategies proposed, we derive valid standard errors and provide software for implementation (R package, *SAMBA*). This paper is the first of its kind to develop a comprehensive statistical framework addressing misclassification and selection bias simultaneously. We provide a new suite of statistical estimation and inference methods tailored to EHR data analysis. Through a simulation study, we demonstrate the ability of these methods to reduce or eliminate bias and correctly estimate standard errors. We apply our proposed methods to address bias (e.g. in GWAS results) and identify factors related to phenotype misclassification using data from The Michigan Genomics Initiative (MGI), a longitudinal EHR-linked biorepository within Michigan Medicine.

## 2 Model, notation, and conceptual framework

Let binary *D* represent a patient’s true disease status for a disease of interest. Suppose we are ideally interested in the relationship between *D* and person-level information, *Z. Z* may contain genetic information, lab results, age, gender, or any other characteristics of interest. We call this relationship the *disease mechanism*.

We consider a large EHR database with the goal of making inference about some defined population. Let *S* indicate whether a given person in the population is included in our data (e.g., by going to a particular hospital and consenting to share biosamples), where the probability of an individual being included in our data may depend on the underlying disease status, *D*, along with additional covariates, *W*. Let *W* ^†^ denote variables in *W* that are not adjusted for in the disease model (not in *Z*). Here, we will use the terms “sampled” or “selected” interchangeably to refer to patients included in our EHR dataset. We may often expect the sampled and non-sampled patients to have different rates of the disease, and other factors such as patient age, residence, access to care, and general health state may also impact inclusion. We will call this mechanism the *selection mechanism*. In reality, patient inclusion in the analytical dataset may be impacted by multiple phases of selection as illustrated for MGI in **Figure C.1**. In our notation, we focus on the aggregate mechanism governing inclusion, which may be composed of these various sub-stages.

Instances of the disease are recorded in the EHR. Factors such as patient age, length of follow-up, and number of hospital visits may impact whether we *observe/record* the disease of interest for a given person. Let *D*^*^ be the *observed* disease status. *D*^*^ is a potentially misclassified version of *D*. We call the mechanism generating *D*^*^ the *observation mechanism*. We will assume that misclassification is primarily through underreporting of disease. In other words, we assume that *D*^*^ has perfect specificity and potentially imperfect sensitivity with respect to *D*. Let *X* denote patient and provider-level predictors related to sensitivity, and let *X*^†^ denote the variables in *X* not included in *Z*. **Figure 1** shows the conceptual model, which is expressed mathematically in *Eq. 1*.

**Figure 1:**
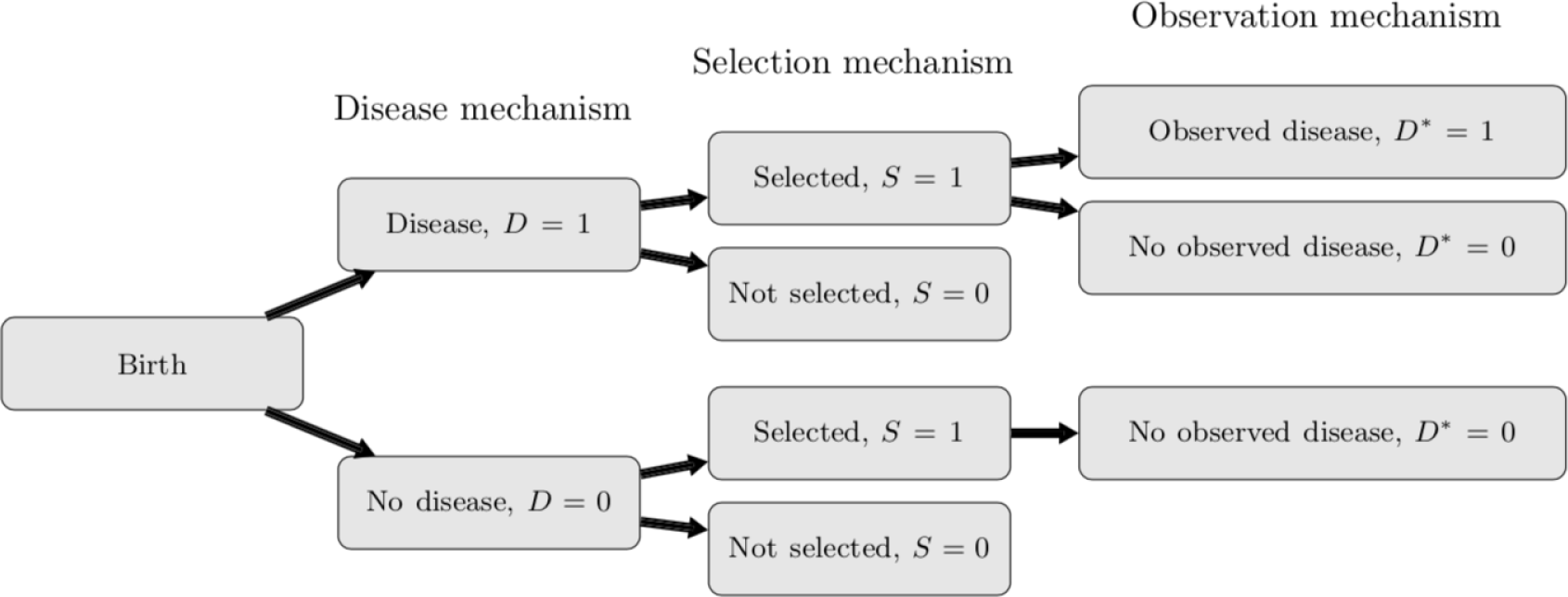
Diagram of the assumed data structure.

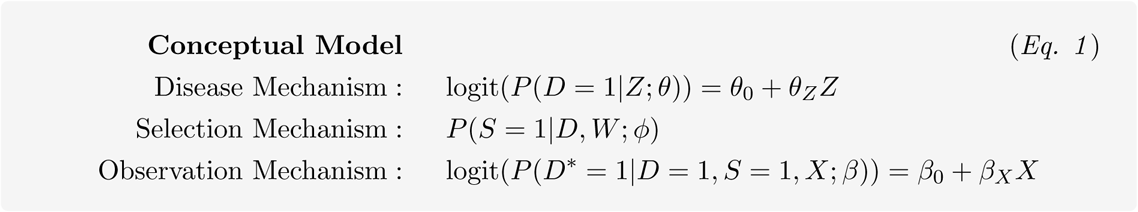

In our statistical development, we will often refer to the following functions of the observation and selection model parameters

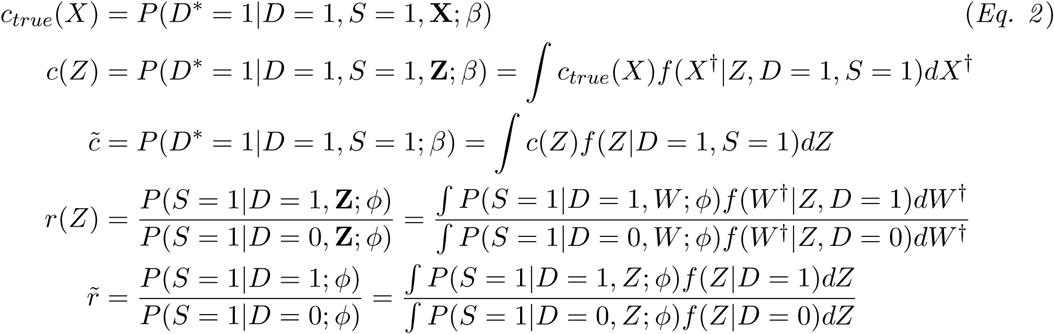

The first expression represents the generating sensitivity mechanism. The subsequent expressions show the average sensitivity as a function of *Z* and the overall marginal sensitivity, 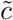, both of which are implicit functions of *β*. The fourth expression represents the sampling ratio with respect to *D* as a function of *Z*, and constant 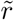 represents the ratio of marginal sampling probabilities (here, called the marginal sampling ratio). These latter expressions are implicit functions of *ϕ*.

A common approach is to model *D*^*^|*Z, S* = 1 (analysis model) and interpret results under the target model *D*|*Z*. There are many settings in which the resulting inference will be biased. To explore these settings, we relate the parameters in the conceptual and analysis models. In **Web Appendix A.1**, we prove the following key relationship:

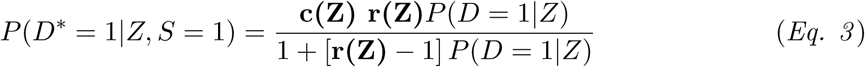

*Eq. 3* is an extension of Neuhaus (1999) allowing for covariate-related misclassification and also incorporating patient selection. The contribution of misclassification and selection reduces to terms *c*(*Z*) and *r*(*Z*) in *Eq. 2*, where *c*(*Z*) represents the impact of misclassification and *r*(*Z*) represents the impact of selection. Under distinctness of *β* and *ϕ* in *Eq. 1, c*(*Z*) and *r*(*Z*) are independent functions of model parameters given *Z*. These two factors work together to generate bias in *P*(*D*^*^ = 1|*Z, S* = 1) relative to *P*(*D* = 1|*Z*). We study settings in which *c*(*Z*) = *r*(*Z*) = 1 in **Web Appendix A.2**. A special case is when *D*|*Z* follows a logistic regression as in *Eq. 1*. In this case, we can show that

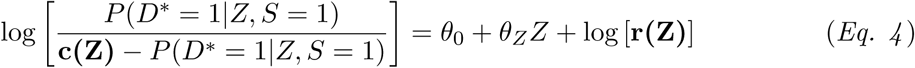

The left-hand side of the equation takes a GLM-type form with a different (non-logistic) non-linear link function, and the right-hand side contains an offset term as a function of the sampling ratio. If we knew both *c*(*Z*) and *r*(*Z*), estimation of *θ* would simply involve fitting the above model to the observed data. In this paper, we provide strategies for estimating *θ* when *c*(*Z*) and *r*(*Z*) are not known, all guided by the relationship in *Eq. 4*.

## 3 Accounting for phenotype misclassification assuming ignorable selection

Suppose first that patient selection is ignorable for *θ*. In other words, assume that *r*(*Z*) = 1. In this case, *Eq. 3* gives that 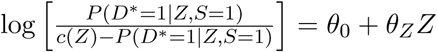. In this section, we propose strategies for estimating *θ* accounting for unknown *c*(*Z*).

### 3.1 Method 1: approximating *D*^*^|*Z*

Suppose *c*(*Z*) is independent of *Z*, so 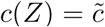. This will be the case if *X* is independent of *Z* given *D*. In this setting, we observe that 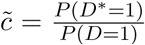. If we know prevalence *P*(*D* = 1), then we can estimate sensitivity as the ratio of observed and true disease prevalences. In Beesley et al. (2018a), we derived an expression relating the true log-odds ratio *θ*_*Z*_ to the uncorrected parameter 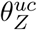 and 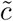 based on Taylor series approximations. As shown in **Web Appendix A.3**, we can further relate *θ*_*Z*_ and 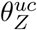 as follows:

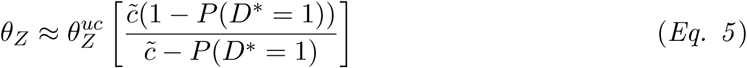

Replacing 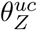 with an estimate, this expression recovers an existing estimator for binary predictor *Z* in Duffy et al. (2004). We show we can apply *Eq. 5* more generally to estimate *θ*_*Z*_ when *Z* and *X* are independent given *D*. This expression is convenient, because it can be applied in settings where only summary statistics for 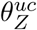 are available.

### 3.2 Method 2: direct estimation of *θ* using a non-logistic link

Suppose instead that *c*(*Z*) is not constant in *Z*. Given *c*(*Z*), we can estimate *θ* using the relationship 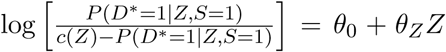, which is a generalized linear model with a non-logistic link function. The question then becomes how to estimate *c*(*Z*).

In **Web Appendix A.4**, we discuss settings where we can replace *c*(*Z*) with estimated *c*_*true*_(*X*) = *P*(*D*^*^ = 1|*D* = 1, *X*; *β*). In practice, replacing *c*(*Z*) with *c*_*true*_(*X*) tends to produce decent results when the covariate of interest is not a direct driver of misclassification. As shown in **Web Appendix A.5**, we can estimate *c*_*true*_(*X*) (function of *β*) using the relationship

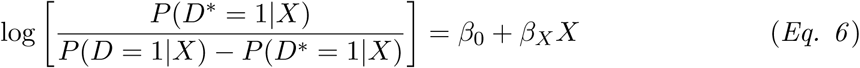

assuming *P*(*D* = 1|*X*) is known. In practice, we will approximate *P*(*D* = 1|*X*) as discussed in **Web Appendix A.5**. Importantly, *Eq. 6* may not always have a solution, and we can instead estimate 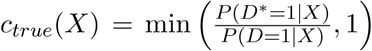. In our experience, this latter approach tends to be more robust to misspecification of *P*(*D* = 1|*X*).

### 3.3 Method 3: joint estimation of *β* and *θ* using observed data log-likelihood

Rather than estimating sensitivity and *θ* in a two-step process, we can *jointly* estimate *θ* and sensitivity parameter *β*. Under assumptions discussed in **Web Appendix A.6**, we estimate (*θ, β*) using the following observed data log-likelihood

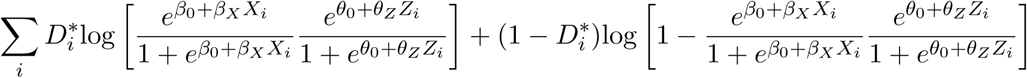

This can be viewed as a zero-inflated logistic regression model as proposed in the context of infection and cure modeling in Diop et al. (2011). This model is identifiable if we have a continuous covariate that is included in *X* but not *Z* or vice-versa. For EHR data, we expect factors such as length of follow-up in the EHR to be included in *X* but not *Z*. We can jointly estimate *θ* and *β* by maximizing this log-likelihood through a Newton-Raphson algorithm or other numerical optimization method. For large datasets, it can sometimes be more computationally convenient to estimate parameters using the expectation-maximization algorithm. In both cases, estimation using the profile likelihood method across *β*_0_ tends to have good performance. We may run into numerical problems tied to weak identifiability in practice, which can be reduced by fixing a model parameter. In simulations, we observed good performance when *β*_0_ was fixed at logit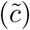 for 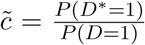 and mean-centered *X*. Estimation details are presented in **Web Appendix A.6**.

## 4 Accounting for patient selection under perfect classification

Suppose that we have some unobserved mechanism governing patient selection and that selection is related to *D*. Additionally, we suppose that we observe *D* perfectly, so *D*^*^ = *D*. In this case, we can relate the observed and true data models using *Eq. 3* as follows: logit [*P*(*D* = 1|*Z, S* = 1)] = *θ*_0_ + *θ*_*Z*_*Z* + log [*r*(*Z*)], where *r*(*Z*) is defined in *Eq. 2*. When *r*(*Z*) is known, we can directly estimate *θ* by fitting the above model. In the setting of case-control sampling, for example, *r*(*Z*) is a (often known) constant and does not impact estimation of *θ*_*Z*_. When *r*(*Z*) is a function of *Z*, however, failure to account for *r*(*Z*) can result in bias. Estimation of *r*(*Z*) directly can be very challenging, and researchers have developed many statistical strategies (e.g. covariate adjustment, propensity weighting, matching, etc.) for obtaining estimates of *θ* without requiring *r*(*Z*). Here, we describe how two such methods can be applied in the EHR setting, and we extend these methods to incorporate selection composed of many intermediate sampling stages.

### 4.1 Method 1: inverse of the selection probability weighting using external data

When sampling probabilities are known or estimable, inverse probability of selection weighting (IPW) can be applied to correct for selection bias. In this approach, we can estimate *θ* by fitting a weighted regression for *D*|*Z* on the sampled data, where each patient’s data is weighted proportional to the inverse of his/her estimated probability of being sampled, *P*(*S* = 1|*D, W*). Estimation of *P*(*S* = 1|*D, W*) for EHR data is generally difficult. However, we can borrow results from the non-probability sampling literature and leverage limited external data from the population of interest to obtain *rough* estimates.

In particular, suppose we have individual-level data on *D* and *W* (or some subset) for an *external* sample of patients from the population of interest. We suppose either the external sampling mechanism or sampling weights is *known*. Example sources of data from the US adult population might include National Health and Nutrition Examination Survey (NHANES); the NCI Surveillance, Epidemiology, and End Results (SEER) program; the CDC Behavioral Risk Factor Surveillance System; and the US Census. We can estimate the selection probability for our *internal* EHR dataset as follows.

Let *S* indicate inclusion in the internal EHR dataset (our analytical dataset), *S*_*ext*_ indicate inclusion in the external data (e.g. NHANES), and *S*_*all*_ indicate inclusion in either the internal or external data. We will assume no one is included in both datasets. Following Elliot (2009) and the proof in **Web Appendix A.7**, we can write

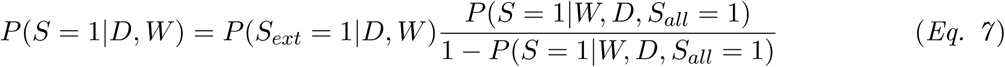

where *P*(*S*_*ext*_ = 1|*D, W*) is the sampling distribution for the external data. When only sampling weights are available, we can estimate *P*(*S*_*ext*_ = 1|*D, W*) by fitting a regression model, e.g. beta regression, for the sampling weights in the external data (Elliot, 2009). We can estimate *P*(*S* = 1|*W, D, S*_*all*_ = 1) by fitting a regression model for inclusion in the internal EHR data given inclusion in the combined dataset. We then define inverse probability of selection weights 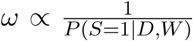, where we approximate *P*(*S* = 1|*D, W*) using available data on *D* and *W* in practice. In many EHR settings, we may want to incorporate more complicated selection steps into the modeling of the aggregate selection mechanism *P*(*S* = 1|*D, W*), as is done in Haneuse and Daniels (2016). In **Web Appendix A.9**, we extend these methods to incorporate multiple selection stages.

### 4.2 Method 2: calibration weighting using external summary statistics

Calibration weighting uses *summary statistics* from the population (e.g., the relationship between *D* and *W*) to re-weight the internal data so that the weighted distributions match distributions in the population. We then estimate *θ* by fitting a weighted regression for *D*|*Z* on the internal sample. Several versions of calibration weighting exist. We will focus on two types: (1) poststratification, where the joint distribution of *D* and *W* (or some subset) is available, and (2) raking, where only marginal distributions are available. Under poststratification, we define weights 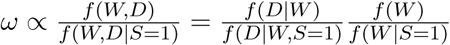, which incorporates summary information from the population along with estimated relationships from the EHR data. We relate these weights to inverse probability of selection weights in **Web Appendix A.7**. Construction of raking weights requires an iterative algorithm to ensure that the resulting weights *ω* recover the marginal distributions in the population. Both weighting strategies can be applied for generalized linear models using R package *survey*.

## 5 Jointly addressing phenotype misclassification and patient selection

When *c*(*Z*) and selection weights *ω* are known, adjustment for both sources of bias is a simple extension of **Section 3** to incorporate weighting. However, sensitivity and weights *ω* will not be known in general, and estimation of these quantities in the presence of both sources of bias is much harder than with each source of bias separately.

Firstly, misclassification complicates the estimation of weights for selection bias adjustment. When we have misclassification, true *D* is not always known, making it difficult to estimate *P*(*S* = 1|*W, D*) or *f* (*D, W* |*S* = 1) directly. Methods in **Section 4** cannot be applied directly. Secondly, sensitivity estimates in **Section 3** often rely on the differences between observed and population disease rates, which will be impacted by the selection mechanism, and cannot be directly applied when we have selection bias. Each source of bias complicates estimation of terms used to correct for the other source of bias, and additional thought is needed to estimate sensitivity and *ω* when both biases are present.

As visualized in **Figure 2**, we propose a series of intermediate steps through which these quantities can be estimated. Fixing these quantities, we then describe how we can estimate *θ* following *Eq. 4*.

**Figure 2:**
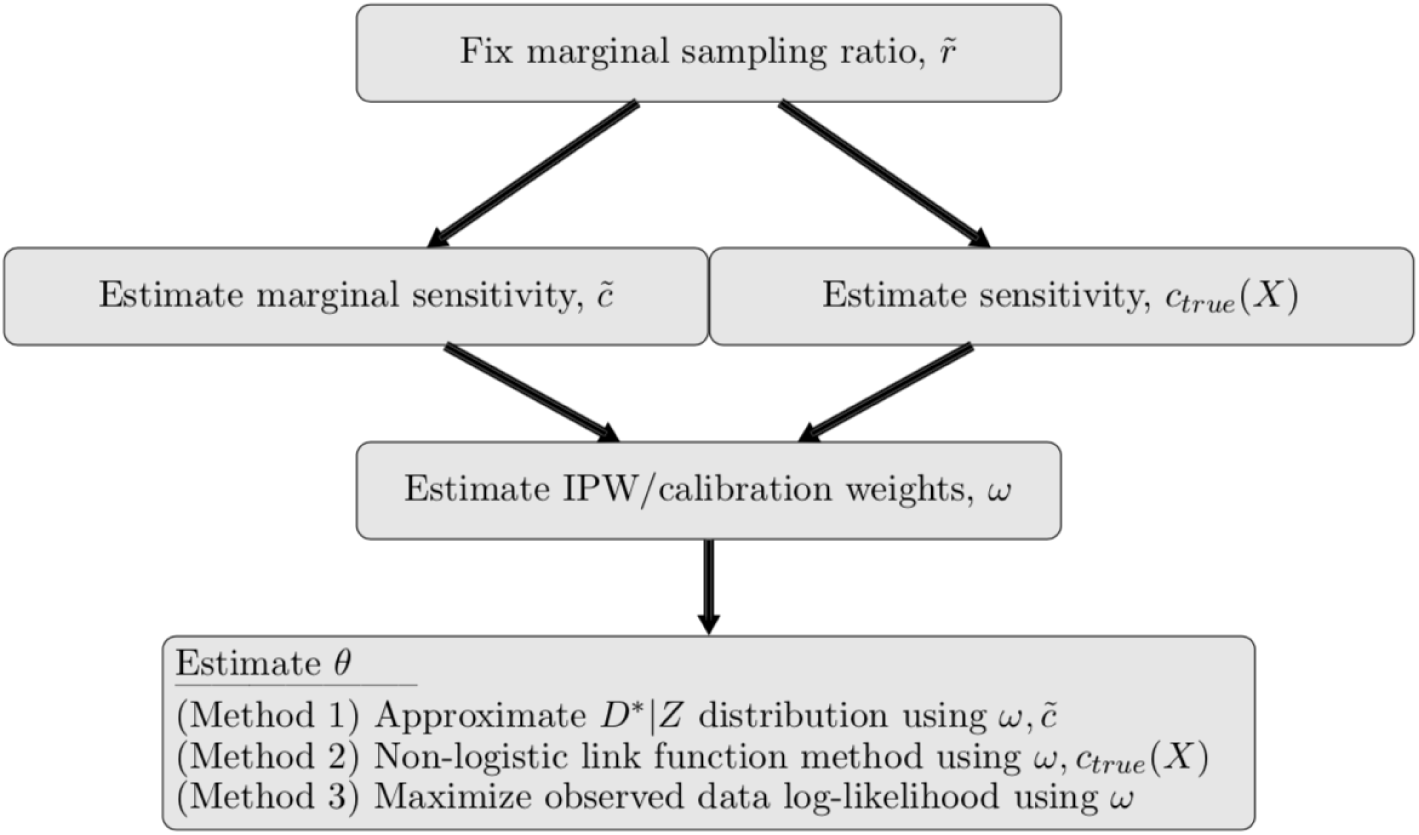
Flowchart of data analysis accounting for both misclassification and patient selection.

### Step 1: Estimating the marginal sampling ratio

In order to estimate *θ*, we first specify the marginal sampling ratio, 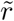. This can be treated as a tuning parameter. We can use the data, the population disease rate *P*(*D* = 1), and our prior beliefs about 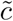 to inform plausible 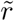 as follows (**Web Appendix A.8**):

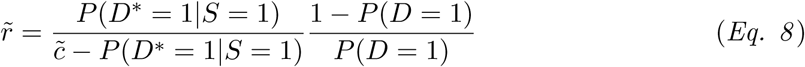

### Step 2: Estimating marginal or patient-specific sensitivities

Given 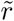, we estimate either marginal sensitivity 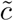 or patient-specific sensitivity *c*_*true*_(*X*). We can estimate 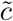 using 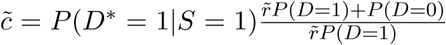, noting that this could produce a sensitivity greater than 1. Instead, suppose we want to estimate *c*_*true*_(*X*). We can estimate *β* (and *c*_*true*_(*X*)) using the following approximate relationship:

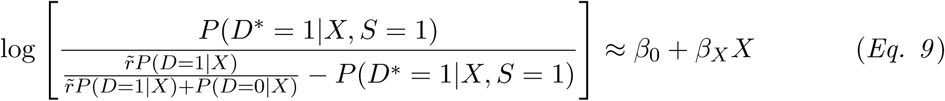

where *P*(*D* = 1|*X*) is specified as in **Web Appendix A.5**. This fit may have no solution for P (D = 1|X) incompatible with the data, and we may directly estimate 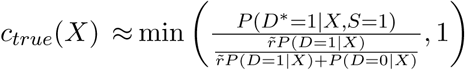 where *P*(*D*^*^ = 1|*X, S* = 1) is estimated using the EHR data.

### Step 3: Estimating sampling or calibration weights

Given 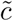 or *c*_*true*_(*X*), we can estimate weights *ω* for selection bias adjustment. When *D* is misclassified, we propose defining inverse probability weights using *P*(*S* = 1|*W, D*^*^) instead of *P*(*S* = 1|*W, D*). Suppose we have individual-level data on *W* (or subset) and *D* for an external sample from the population. Approximating *P*(*S* = 1|*W, D*^*^) with *P*(*S* = 1|*W, D*^*^) (after replacing *W* with available subset), we estimate the selection probability using (**Web Appendix A.7**):

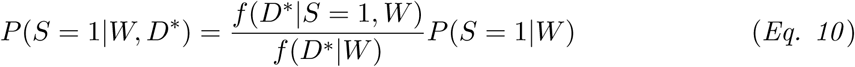

Each element of *Eq. 10* is estimable using the external data and internal EHR. *P*(*S* = 1|*W*) does not involve *D*, so it can be directly estimated using *Eq. 7* or methods in **Section A.9**. *P*(*D*^*^ = 1|*S* = 1, *W*) can be directly estimated using the internal data. Combining sensitivity *c* (estimated using 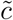 or *c*_*true*_(*X*)) with estimated *P*(*D* = 1|*W*) from the external data, we can express *f* (*D*^*^|*W*) ≈ [*cP* (*D* = 1|*W*)]^*D*\*^ [1 - *cP* (*D* = 1|*W*)]^1−*D*\*^. Using similar logic, we can define poststratification weights as 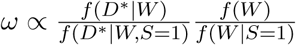.

### Step 4: Estimating *θ* given sampling/calibration weights *ω* and sensitivity

### 5.1 Method 1: approximation of *D*^*^|*X* accounting for selection

Suppose we assume *r*(*Z*) is a constant. This may be reasonable if *Z* is independent of *W* ^†^ given *D* and the covariates of interest in *Z* are not contained in *W*. Suppose further that *c*(*Z*) can be replaced with constant 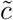. Then, we can apply *Eq. 5* directly to correct for both sources of bias (**Web Appendix A.3**). Intuitively, the impact of the selection enters that estimator through the observed rate of disease in the sample. In general, *r*(*Z*) may rarely be a constant, and we show that 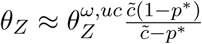, where 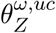 is estimated from fitting a model for *D*^*^|*Z* on the *sampled* patients and *weighting* by *ω*, and *p*^*^ is the *ω*−*weighted* average of *D^*^* in our sample (**Web Appendix A.3**).

### 5.2 Method 2: non-logistic link function method with weighting

Suppose sensitivity is a function of *Z*. Recall from *Eq. 4* that 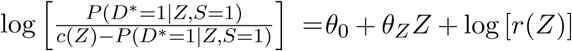. We again remove the contribution of *r*(*Z*) by weighting estimation by *ω*. In particular, we estimate *θ* by fitting an *ω*-weighted version of the model 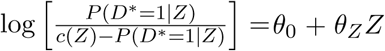 to the patients in the sample. Assuming *X*^†^ is independent of *Z* given *D*, we can replace *c*(*Z*) with estimated *c*_*true*_(*X*).

### 5.3 Method 3: joint estimation using observed data log-likelihood incorporating weights

We can jointly estimate *θ* and *β* accounting for selection bias by maximizing a *ω*−weighted version of the log-likelihood in **Section 3.3** or applying a weighted expectation-maximization algorithm. Details are available in **Web Appendix A.6**.

## 6 Standard error estimation for bias-corrected estimates

Until this point, we have focused on point estimation for *θ*, but we are also interested in valid standard errors. In **Web Appendix A.10**, we detail how to estimate standard errors for each of the proposed methods. Here, we summarize that discussion. When we do not account for selection bias, variance estimation is more straightforward. For the method in **Section 3.1**, we can estimate standard errors for 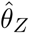 given 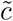 as 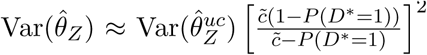. When we estimate *θ* given *c*(*Z*) as in **Section 3.2**, the corresponding information matrix can be inverted to obtain standard errors for 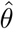. Similarly, we can obtain an estimated covariance matrix after joint estimation of *θ* and *β* from **Section 3.3** using the inverse of the expected observed data information matrix. We analytically show that our bias correction methods ignoring selection will result in *larger* standard errors relative to naive analysis on average (**Web Appendix A.10**). Joint estimation of *θ* and *β* will produce the largest standard errors on average.

Methods for selection bias adjustment ignoring misclassification involve fitting a weighted regression model. Corresponding standard errors can be estimated using a Huber-White sandwich estimator as implemented in the R package *survey* (Freedman, 2006). We can obtain standard errors for weighted versions of the first two *θ* estimation methods in **Section 5** similarly. In order to estimate standard errors for the observed data log-likelihood method with weighting, we propose a sandwich estimator based on weighting the observed data score and information matrices as detailed in **Web Appendix A.6**.

For the majority of these methods, standard errors are estimated fixing sensitivity and/or selection bias weights *ω*. However, rigorous estimation of standard errors should also incorporate uncertainty from estimating these quantities. To account for this residual uncertainty, we could apply bootstrap methods, where the sensitivity, weights, and *θ* are estimated on each of many bootstrap samples of the data. The resulting distribution of 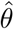 can then be used to estimate standard errors. We compare the degree of underestimation due to ignoring this uncertainty in our simulations. We find that ignoring uncertainty in estimating sensitivity does not impact variance estimation too much, but there is some underestimation of variance when we ignore uncertainty in estimating selection weights.

## 7 Simulations

We present a simulation study to evaluate the proposed methods in terms of bias and bias-corrected inference in settings with disease status misclassification and/or non-ignorable patient selection. We divide this simulation study into three parts. In the first part, we focus on the setting where the outcome is misclassified but patient selection can be ignored. In the second part, we focus on selection and assume we have no misclassification. After evaluating these two simpler cases, we then explore the more complicated setting where we have both sources of bias.

In all simulation settings, we generate 500 datasets with 5000 population members each under a ∼10% marginal disease prevalence. In part 1, we impose outcome misclassification under different covariate-related sensitivity mechanisms corresponding to marginal sensitivities of roughly 0.4, 0.65, 0.8, and 0.95. In part 2, we sub-sample about 50% of patients under various sampling mechanisms. In part 3, we sub-sample patients under different mechanisms *and* impose roughly 65% outcome sensitivity. In each simulation setting, we apply methods discussed in this paper to correct bias in disease model parameters. Details about data generation and implementation can be found in **Web Appendix B.1**.

### 7.1 Simulation results

**Figure 3** presents the biases in the estimated log-odds ratio of *Z* (from *D*|*Z* model, truth = 0.5) across 500 simulated datasets for the first two simulation scenarios. **Figure 4** presents the bias for the third simulation scenario.

**Figure 3:**
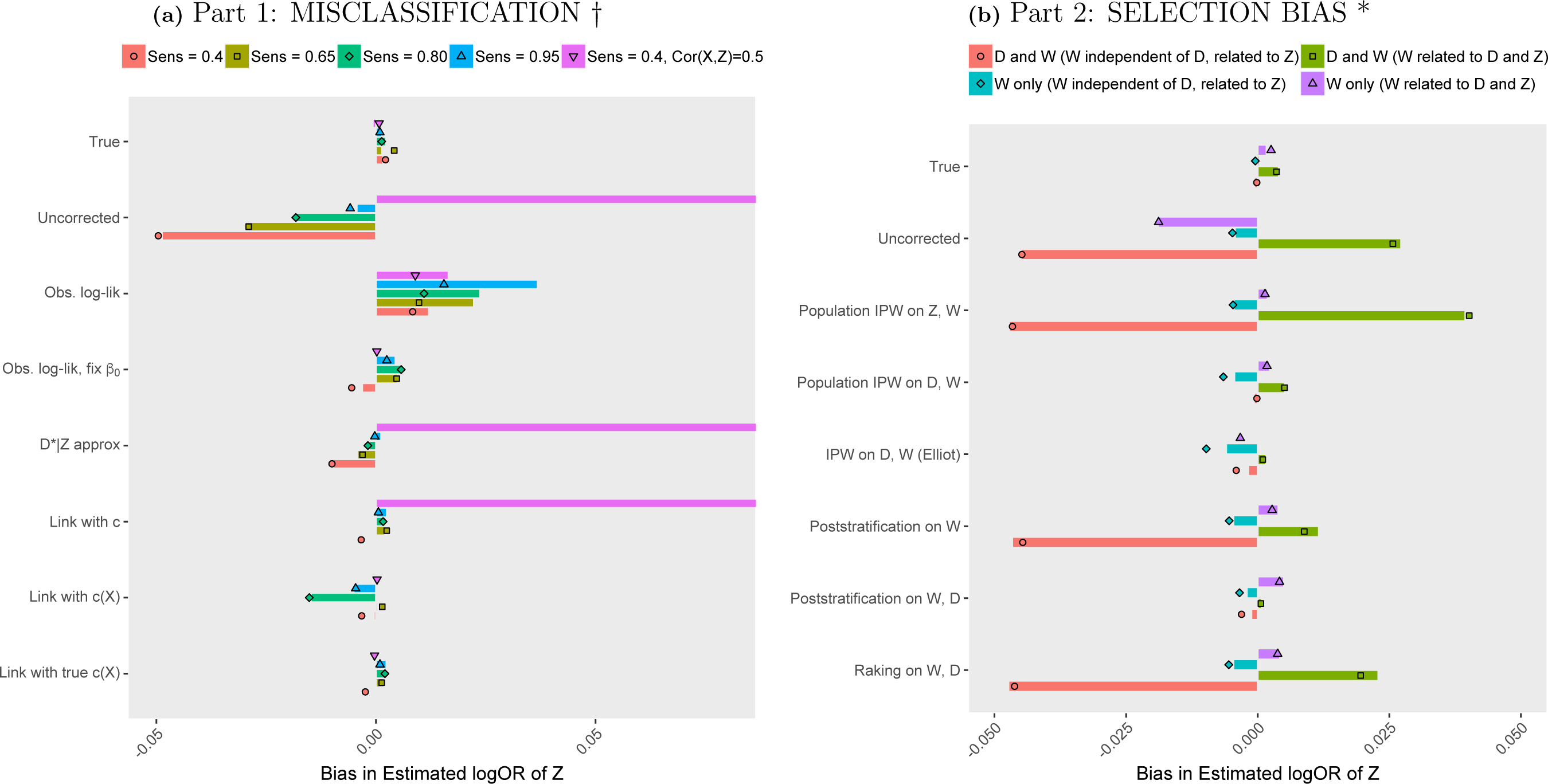
Bias in estimated log-odds ratio of *Z* across 500 simulations under selection bias *or* phenotype misclassification. Bars (points) represent the average (median) difference between estimates and the truth of *θ*_*Z*_ = 0.5. †Here, *c*(*X*) denotes an estimate of *c*_*true*_(*X*) and *c* denotes an estimate of 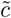. ‘Link’ represents the non-logistic GLM fit. \**Population* IPW indicates the selection model was estimated using data from the entire population.

**Figure 4:**
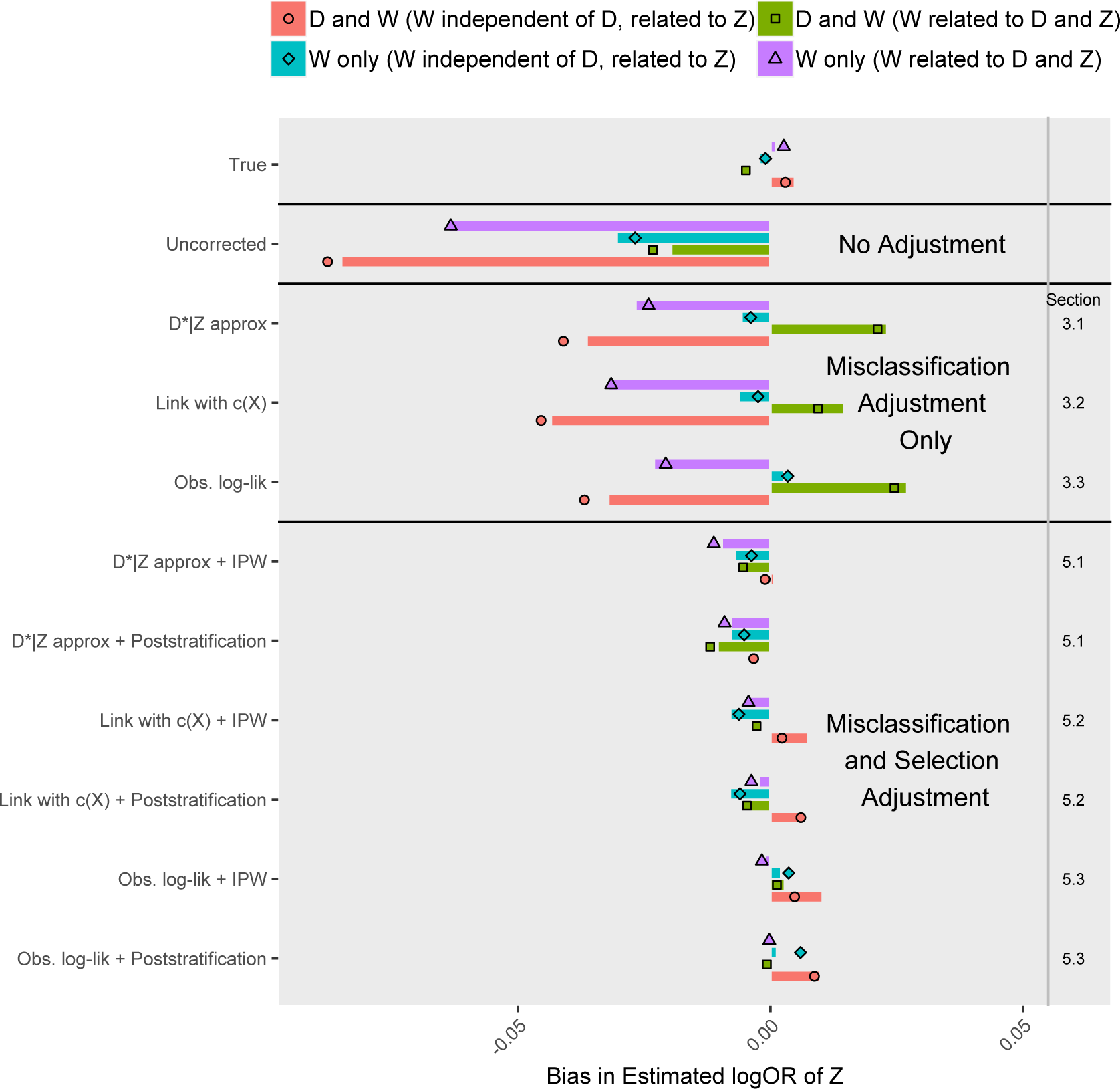
(Part 3) Bias in estimated log-odds ratio of *Z* across 500 simulations under selection bias *and* phenotype misclassification. †** Bars (points) represent the average (median) difference between estimates and the truth of *θ*_*Z*_ = 0.5. †Here, *c*(*X*) denotes an estimate of *c*_*true*_(*X*) and *c* denotes an estimate of 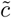. ‘Link’ represents the non-logistic GLM fit. ** IPW was implemented using the *true* selection probabilities. We obtain similar results using *estimated* probabilities.

#### Misclassification Only

Suppose first that *X* and *Z* are independent. Relative bias in uncorrected analysis ranges from very small (high sensitivity) to about 10% (low sensitivity). When *X* ⊥ *Z*, the proposed methods do a good job at removing or reducing bias to less than 2%. An exception is the observed data log-likelihood maximization method, which maintains residual bias due to numerical instability. When we fix *β*_0_ (the intercept in the misclassification model) at a reasonable value, however, this method has excellent performance in terms of bias. When *X* and *Z* are correlated, the uncorrected analysis bias increases to over 40%, and this bias is in the direction away from the null. Methods treating sensitivity as fixed do not correct this bias, but methods that treat sensitivity as a function of covariates reduce this bias to much smaller levels.

#### Selection Only

Analysis of selected patients without correction produces bias from 4% to 9% except in the setting where *W* is independent of *D* and is the only driver of selection (no bias). These biases are not too large, but they can grow larger with stronger covariate effects on selection. We compare weighting strategies for correcting this selection bias. When the IPW model is correctly structured, we can estimate parameters with low bias. This is true even when the true selection probabilities are not known and must be estimated as in *Eq. 7*. Poststratification on both *W* and *D* using external summary statistics has good performance in terms of bias. When selection is independent of *D* given *W*, poststratification on *W* also performs well. Raking (using marginal distributions of *D* and *W*), however, performs poorly when selection depends directly on *D*.

#### Both Selection and Misclassification

Biases of uncorrected analysis range from about 4% to 17% relative to the true log-odd ratio of 0.5. We see even higher bias if *X* is related to *Z* (e.g. 20%+). Notably, methods that only correct for misclassification can still result in substantial residual bias (3-9%), and this can even be larger than bias in the naïve analysis. When we also adjust for selection, however, we see little bias.

#### Other Metrics for Inference

**Figure 5** provides empirical and estimated variances. The estimated variances tend to be similar or slightly smaller than empirical variances on average. Ignoring uncertainty due to estimation of selection weights seems to be a bigger problem than ignoring uncertainty due to estimation of sensitivity. Coverage rates of 95% confidence intervals tend to be low (as low as 8% in our simulations) for naive analyses. In contrast, coverages tend to be near nominal for methods that fully correct the bias. Simulations in **Web Appendices B.2-B.4** demonstrate that the proposed estimators for sensitivity and *ω* generally do a good job at recovering the true values when models are correctly specified. In **Web Appendix B.2**, we demonstrate that misclassification bias-adjusted p-values tend to be similar to unadjusted p-values when *Z* and *X* are independent. However, when *Z* and *X* are associated, the corrected and uncorrected p-values differ, and uncorrected type I error can be highly inflated (e.g. 0.60).

**Figure 5:**
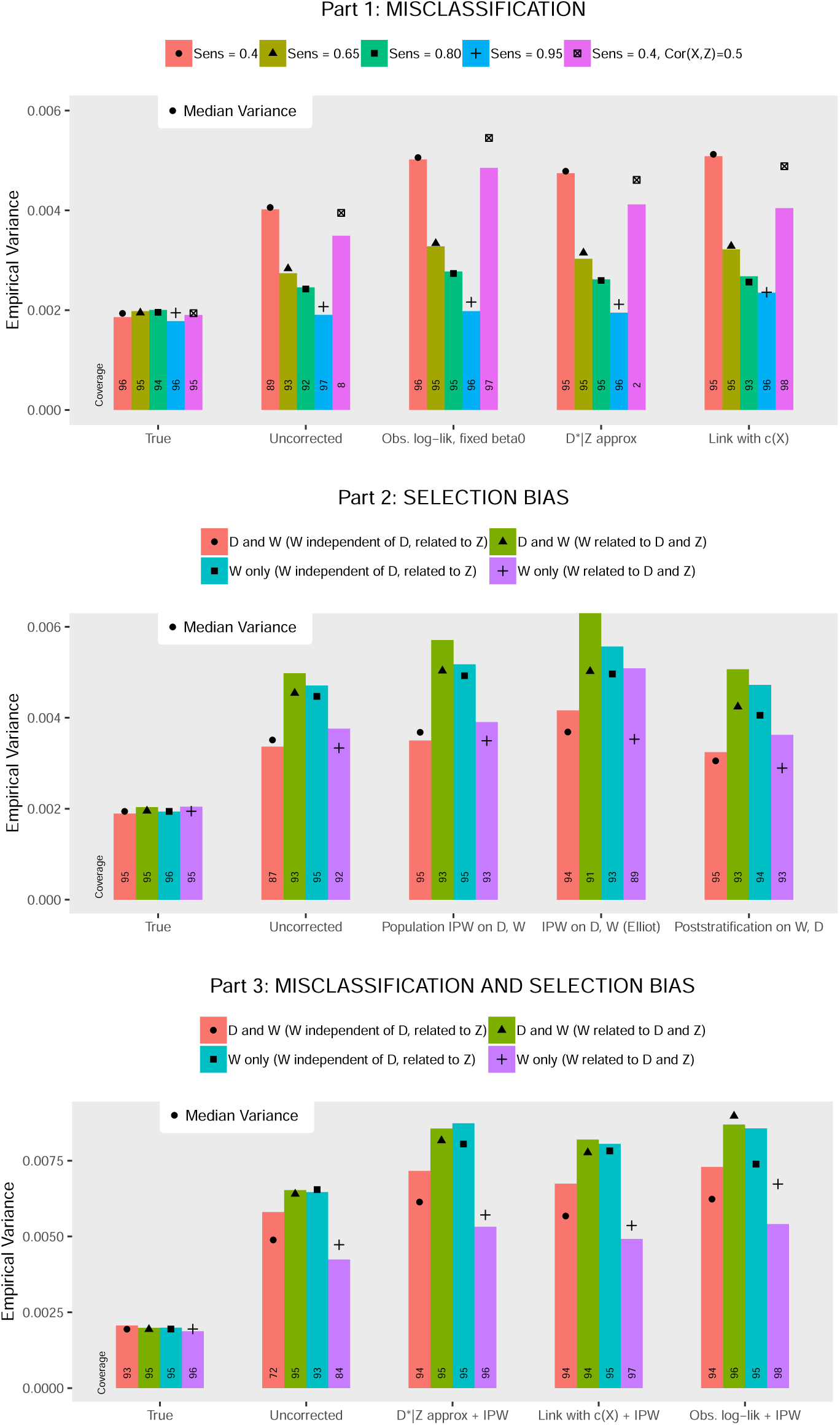
Comparison of empirical and median estimated variances for the log-odds ratio of Z across 500 simulations^†^. † This figure presents empirical variances (bars) and median estimated variances (points) across 500 simulated datasets. We calculate these quantities after applying a variety of bias-correction strategies. Here, *c*(*X*) denotes an estimate of *c*_*true*_(*X*). ‘Link’ represents the non-logistic GLM fit. When handling selection only, *Population IPW* indicates the selection model was estimated using data from the entire population, and *Elliot* indicates the selection model was estimated as in *Eq. 7*. When handling misclassification *and* selection, true selection probabilities were used. Coverage of 95% confidence intervals is printed at the bottom of each bar.

## 8 Data example: exploring bias and sensitivity in MGI

The Michigan Genomics Initiative (MGI) is an EHR-linked biorepository within Michigan Medicine containing over 40,000 patients with matched genotype and phenotype information (Fritsche et al., 2018). **Table C.1** provides descriptives of the unrelated patients of recent European ancestry used for our analyses. Time-stamped ICD (International Classification of Disease) diagnosis data are available for each patient. We map ICD codes to a set of 1866 diseases and symptoms called “phenotype codes” or “phecodes” (Carroll et al., 2014). Observed disease status, denoted *D*^*^, indicates whether each patient received a given phecode during follow-up. Let *D* denote the “true” disease status.

**Figure C.1** provides a visualization of the MGI data accumulation process. We are concerned about generalizing results in MGI to external populations. Patients in a hospital EHR will naturally be sicker than the average person in the US. Moreover, MGI, a subset of Michigan Medicine, primarily recruits perioperative patients, resulting in strong enrichment for many diseases. When ignored, these factors could result in a large degree of bias in association analyses. We apply the proposed methods to address phenotype misclassification and selection bias in MGI through three illustrative examples. In the first example, we explore factors measured in the EHR that may be related to misclassification for several diseases of interest. In the second example, we apply the proposed methods to study a well-known relationship between gender and cancer risk. In the third example, we examine bias in genetic association study results for age-related macular degeneration. **Table C.2** clarifies the link between each example and the conceptual framework laid out in *Eq. 1*.

### 8.1 Example 1: factors related to sensitivity in MGI

The proposed methods in **Section 3** provide a unique opportunity to explore factors related to sensitivity in EHR data. We study five EHR-derived phenotypes in MGI: cancer, colorectal cancer, diabetes, hypothyroidism, and melanoma. We modeled sensitivity as a function of follow-up years, age, and the number of doctor’s visits per follow-up year. We adjusted for gender and age in the disease model. Results are shown in **Figure 6**.

**Figure 6:**
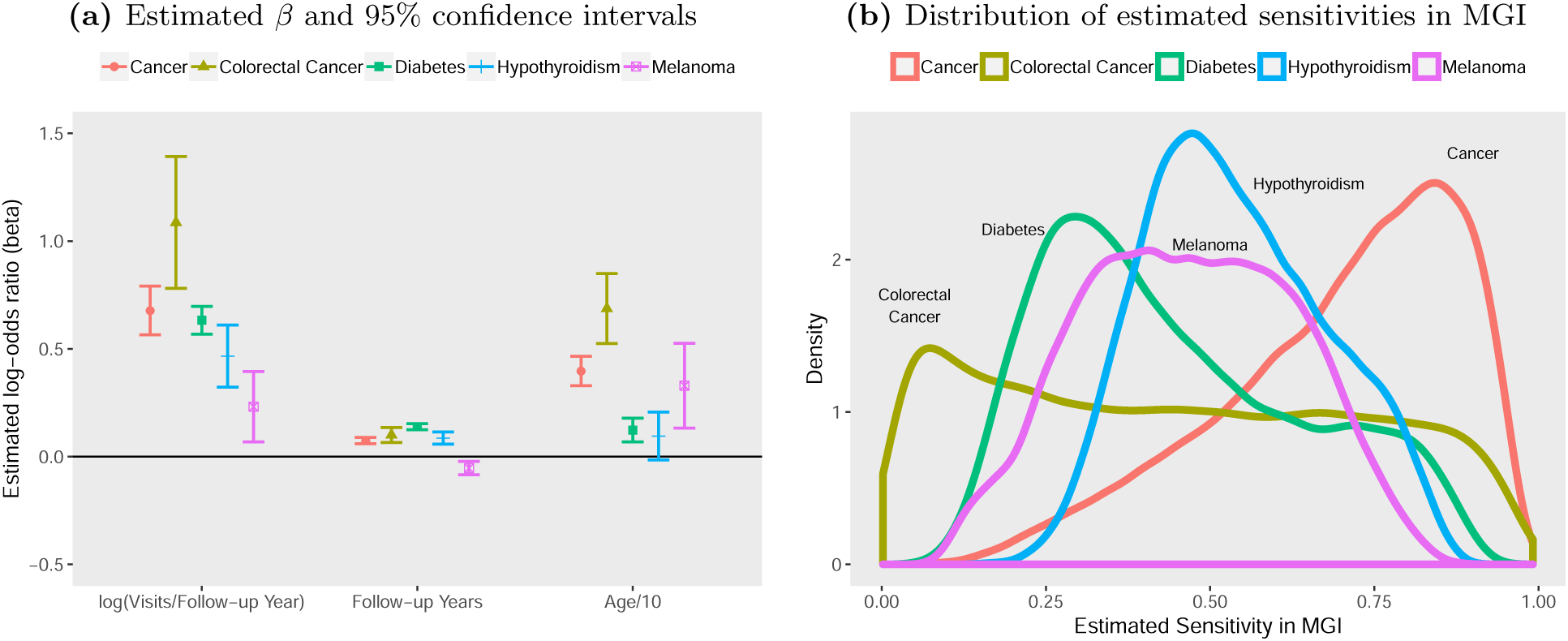
Sensitivity of EHR-derived disease phenotypes in MGI*. *Sensitivity was estimated using the method in **Section 3.3** ignoring selection and adjusting for age and gender in the disease model.

**Figure 7:**
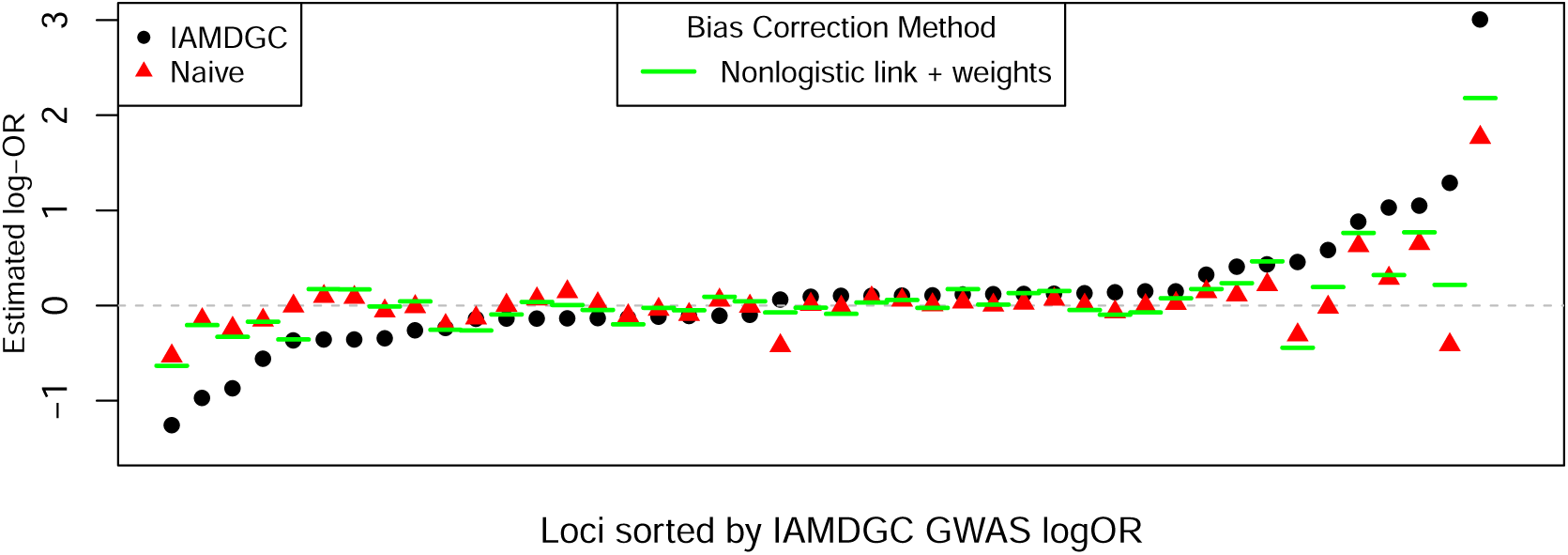
AMD log-odds ratio estimates across 44 genetic loci in MGI and IAMDGC*. *We show log-odds ratio estimates for 44 genetic loci known to be associated with AMD. Circles correspond to the results from the IAMDGC GWAS, here viewed as a comparative gold standard. Triangles show estimates in MGI ignoring selection and misclassification-related biases. Horizontal lines correspond to estimated associations in MGI after applying the method in **Section 5.2**.

Average estimated sensitivity across patients ranged from 0.40 for diabetes to 0.73 for cancer, but individual patients’ sensitivity estimates differed greatly. This supports a need to consider covariate relationships with sensitivity in EHR data. A greater number of visits per follow-up year was associated with higher sensitivity, with odds ratios associated with log(visits per year) ranging from 1.26 (95% CI: 1.07, 1.48) for melanoma to 2.96 (95% CI: 2.18, 4.02) for colorectal cancer. The odds ratio for years of follow-up was greater than 1 for all diseases except melanoma (0.95 [95% CI: 0.92, 0.98]). Michigan Medicine is well-known for its skin cancer clinic, and many patients receive treatment over a short period of time and then return to their home clinics for subsequent care. This could explain an inverse relationship between follow-up duration and accuracy of EHR-derived skin cancer diagnosis. Increased age was associated with higher sensitivity for all diseases considered, with odds ratios up to 1.99 (95% CI: 1.69, 2.24) for colorectal cancer.

### 8.2 Example 2: assessing association between gender and cancer risk

Suppose we are interested in the relationship between cancer (*D*) and gender (*Z*). Odds ratios reported using SEER data indicate lower cancer risk among women, with estimates of 0.78 (2008-2010), 0.83 (2010-2012), 0.92 (2012-2014), and 0.94 (2014-2016). An estimate using 28,709 patients in NHANES (2011-2016) is 0.88 (95% CI: 0.77, 0.99).

We first present an illustrative example (example 2a) where we treat EHR-derived cancer status as the truth and impose additional misclassification. We then present example 2b, where we correct for an *unknown* degree of misclassification in EHR-derived cancer status along with potential selection bias using the methods in **Section 5**.

Example 2a: Suppose we treat EHR-derived cancer diagnosis as the *truth, D*. In this case, we estimate the “true” gender odds ratio in MGI as 0.93 (95% CI: 0.90, 0.97), where the reference category is men. Given *D*, we then impose misclassification (generate *D*^*^) using two different mechanisms: (1) patients with longer follow-up are more likely to have observed disease and (2) patients with longer follow-up and female patients are more likely to have observed disease, each resulting in an average sensitivity of about 70%. We apply methods in **Section 3** to correct resulting bias in the gender odds ratio.

**Figure C.2** shows the associations between gender and the misclassified outcome. In both settings, some bias is evident and is particularly notable when misclassification depends directly on gender (odds ratio 1.01 [95% CI: 0.97, 1.06]). When misclassification does not depend directly on gender, our methods can correct the bias even when the sensitivity model parameters are estimated. When misclassification depends directly on gender, however, it is more difficult to estimate sensitivity, and our methods struggle unless the true sensitivity is known. In general, our methods have good performance in correcting bias when sensitivity is not strongly related to the predictor of interest, *Z*.

<u>Example 2b:</u> Now, we apply the proposed methods to correct bias already present in the observed MGI data. Suppose our population of interest is the general US population, and we define the *true* cancer status of interest, *D*, as the patient’s historical cancer status up to their current age. We now want to study the relationship between *true* cancer status *D* and gender (*Z*), adjusting for potential misclassification of the EHR-based cancer diagnosis status (*D*^*^) and the unknown mechanism driving patient inclusion in the EHR.

We estimate sensitivity given age, length of follow-up, and log(number of visits per follow-up year) using *Eq. 9* and fixing unknown 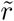 at 250, resulting in a median estimated sensitivity of roughly 0.64 and ranging from 0.11 to 0.96 across patients. We adjust for selection bias using poststratification and inverse probability of selection weighting using several weight construction strategies. Details can be found in **Web Appendix C.4**.

We apply methods described in **Sections 3 and 5** to estimate the association between cancer and gender. Results are shown in **Table 1**. First, we adjust for misclassification only. Using the non-logistic link function and observed data log-likelihood methods, we obtain odds ratio estimates of 0.85 (95% CI: 0.75, 0.95) and 0.97 (95% CI: 0.87, 1.08) respectively. When we adjust for both misclassification and selection bias and incorporate cancer diagnosis in the selection weights, point estimates range from 0.77 (95% CI: 0.72, 0.82) to 0.86 (95% CI: 0.77, 0.96). Since our patient population is a mix of diagnoses across these time periods, it is difficult to assess the “truth,” but this exploration does demonstrate the potential impact of bias correction.

**Table 1:**
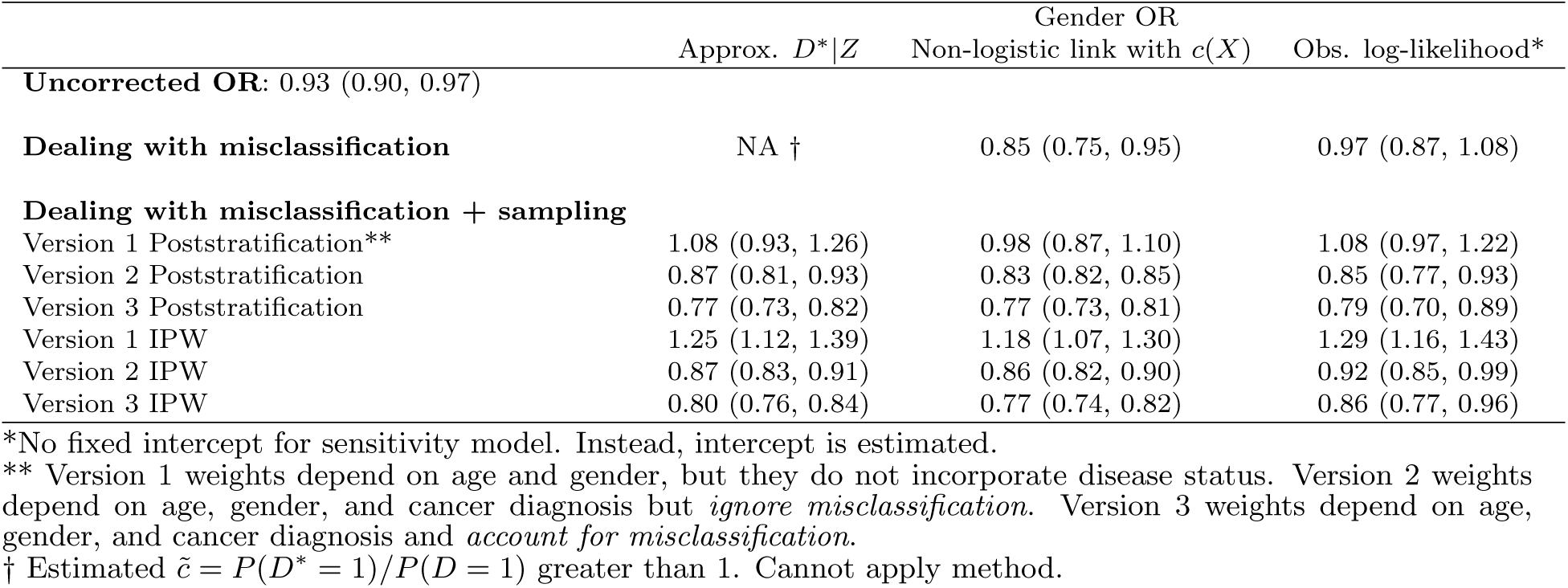
Estimated cancer-gender odds ratio and 95% confidence intervals across methods (reference category = male)

### 8.3 Example 3: genetic associations for age-related macular degeneration

Age-related macular degeneration (AMD) is a common cause of vision loss, and the International AMD Genomics Consortium (IAMDGC) was developed to better understand its genetic drivers. In this section, we compare results from a genome-wide association study of over 16,000 advanced AMD cases and 18,000 controls using IAMDGC data to parallel results using matched MGI case-control data among patients over 50. Details of the data curation are available in **Web Appendix C.5**. We focus on 44 independent genetic loci identified with small p-values (*<*5×10^−8^) in the IAMDGC data. Across these 44 loci, MGI and IAMDGC GWAS log-odds ratio point estimates have a Lin’s concordance correlation coefficient (CCC) of only 0.61, and MGI point estimates generally tend to be closer to the null compared to the IAMDGC estimates (**Figure 5.2**). The “winner’s curse” resulting in inflated IAMDGC point estimates explains some differences, but bias due to selection and misclassification in MGI may also contribute. Another explanation for smaller effect estimates in MGI is that less advanced AMD cases were also included, as were cases of macular degeneration in older adults that may not have been age-related. We apply our methods to correct for potential biases due to selection and misclassification in MGI.

AMD sensitivity in MGI is estimated as a function of age, length of follow-up in the EHR, and log(number of visits per follow-up year). Fixing 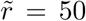 (see **Web Appendix C.5** for details), we obtain a median estimated sensitivity of 0.17 that ranges between 0.01 and 0.99 across patients. We use these sensitivities and poststratification weights obtained using US Census and NIH National Eye Institute summary statistics to correct for potential bias in the MGI estimates. We compare the resulting point estimates in **Table C.4**. When we accounted for both potential selection bias and misclassification, Lin’s concordance increased from 0.61 to 0.77. Additionally, the average absolute difference between MGI and IAMDGC estimates went from 0.21 for naive analysis to 0.13 for bias-adjusted analysis. **Figure 5.2** provides the point estimates in MGI after applying the method in **Section 5.2**. These estimates tend to be closer to IAMDGC estimates than seen in naïve analysis. These results indicate that the proposed bias-correction methods allowed us to better recover genetic associations observed using better quality data.

## 9 Discussion

Electronic health records (EHR) have become a rich resource for biomedical research. However, data obtained from EHR are highly susceptible to various sources of bias, which can negatively impact the accuracy and generalizability of statistical inference. In this paper, we focus on two common sources of bias: (1) misclassification of derived disease variables (information bias), which may be related to patient factors, and (2) lack of representativeness (selection bias). To address these key problems, we propose a variety of likelihood-based bias-correction strategies. We also derive valid standard errors, allowing for principled inference, and we provide corresponding software (R package, *SAMBA*, available at https://github.com/umich-cphds/SAMBA).

A key advancement in this paper is the development of strategies to handle covariate-related phenotype misclassification. The proposed methods leverage information in each patient’s follow-up history to estimate the rate of misclassification *without requiring gold standard disease status labels*. We also describe how we can incorporate external disease information into the estimation, resulting in efficiency gains.

In addition to addressing misclassification, we explore strategies for dealing with the harder problem of selection bias. Correction for selection bias is extremely difficult for EHR data, and we describe how we can extend weighting methods in the survey sampling literature to at least partially address patient selection. As in Haneuse and Daniels (2016), our methods can accommodate the complicated multi-stage selection mechanisms often present for EHR data, but our methods further bridge the gap between patients that are and are not included in the EHR.

The key limitation of these methods is the need for high-quality external information, including external summary statistics or even individual-level data from the population of interest.

This paper proposes a variety of bias-correction strategies. Among the methods for handling misclassification, the method in **Section 3.2** is particularly attractive and easy to implement. Estimating sensitivity under that method requires some external summary information (e.g. association between disease and age, gender), but this may be easily obtained for many diseases. Poststratification emerges as an appealing approach for handling selection bias since it relies on summary statistics from the population rather than individual-level data. The combination of these two approaches in **Section 5.2** tends to produce good results for addressing both sources of bias at once. We recommend this approach as a starting point for analysts interested in applying the proposed methods.

Our results rely on logistic regression models for misclassification and disease, and we assume perfect specificity. Future work should explore more general modeling settings. We focus our attention on a single disease *D* and adjustment factors *Z*, but these methods could be applied to study many disease-covariate combinations. Strategies for automating estimation for association studies are discussed in **Web Appendix D.2**. In general, this paper provides useful statistical strategies and corresponding software for handling outcome misclassification and selection bias, and these methods are tailored to issues encountered in EHR data analysis.

## Data Availability

MGI data are available after IRB approval to select researchers.

## Acknowledgments

The authors would like to thank Chad Brummet, Goncalo Abecasis, and Sachin Kheterpal along with the large group of collaborators and staff at Michigan Genomics Initiative along with MGI participants for generously donating their biosamples for research. This work was supported by The University of Michigan Comprehensive Cancer Center core grant supplement 5P30-CA-046592, NSF DMS award 1712933 and The University of Michigan precision health award U067541. The authors acknowledge the University of Michigan Medical School Central Biorepository for providing biospecimen storage, management, and distribution services in support of the research reported in this publication. We want to thank Lars Fritsche and the International AMD Genomics Consortium (IAMDGC) for providing GWAS summary statistics. We also want to thank Alexander Rix for his help in developing the R package.

## Supporting Information

R package *SAMBA* can be found at https://github.com/umich-cphds/SAMBA. MGI data are available after IRB approval to select researchers.

